# Structural and functional connectomes in people with multiple sclerosis

**DOI:** 10.1101/2020.08.25.20181727

**Authors:** Ceren Tozlu, Keith Jamison, Susan A. Gauthier, Amy Kuceyeski

## Abstract

One of the challenges in multiple sclerosis is that lesion volume does not correlate with symptom severity. Advanced techniques such as diffusion and functional MRI allow imaging of the brain’s connectivity networks, which may provide better insight as to brain-behavior relationships in impairment and compensation in multiple sclerosis. We aim to build machine learning models based on structural and functional connectomes to classify a) healthy controls versus people with multiple sclerosis and b) impaired versus not impaired people with multiple sclerosis. We also aim to identify the most important imaging modality for both classification tasks, and, finally, to investigate which brain regions’ connectome measures contribute most to the classification. Fifteen healthy controls (age=43.6± 8.6, 53% female) and 76 people with multiple sclerosis (age: 45.2 ± 11.4 years, 65% female, disease duration: 12.2 ± 7.2 years) were included. Twenty-three people with multiple sclerosis were considered impaired, with an Expanded Disability Status Scale of 2 or higher. Subjects underwent MRI scans that included anatomical, diffusion and resting-state functional MRI. Random Forest models were constructed using structural and static/dynamic functional connectome measures independently; single modality models were then combined for an ensemble prediction. The accuracy of the models was assessed by the area under the receiver operating curve. Models that included structural connectomes significantly outperformed others when classifying healthy controls and people with multiple sclerosis, having a median accuracy of 0.86 (p-value<0.05, corrected). Models that included dynamic functional connectome metrics significantly outperformed others when distinguishing people with multiple sclerosis by impairment level, having a median accuracy of 0.63 (p-value<0.05, corrected). Structural connectivity between subcortical, somatomotor and visual networks were most damaged by multiple sclerosis. For the classification of patients with multiple sclerosis into impairment severity groups, the most discriminatory metric was dwell time in a dynamic functional connectome state characterized by strong connectivity between and among somatomotor and visual networks. These results suggest that damage to the structural connectome, particularly in the subcortical, visual and somatomotor networks, is a hallmark of multiple sclerosis, and, furthermore, that increased functional coordination between these same regions may be related to severity of motor disability in multiple sclerosis. The use of multi-modal connectome imaging has the potential to shed light on mechanisms of disease and compensation in multiple sclerosis, thus enabling more accurate prognoses and possibly the development of novel therapeutics.

## INTRODUCTION

Multiple Sclerosis (MS) is a chronic disease characterized by inflammatory and demyelinating plaques within the central nervous system (Weinshenker et al., 1991). One key observation is that the disease burden in the brain, as measured with conventional imaging, is not always proportional to an individual’s disability. Therefore, individuals can have identical lesion volume and very different levels of impairment, i.e. the clinic-radiological paradox in MS (Barkhof, 2002). More advanced neuroimaging techniques may enable us to better understand the neuropathological mechanisms of MS, how they cause impairment and how the brain may compensate for this pathology. Brain connectivity network analysis, or connectomics, provide a promising tool to map the effect of MS-related pathology and to potentially capture reorganization mechanisms in response to pathology. The inflammation, demyelination, and axonal loss in people with MS (pwMS) disrupts the brain’s structural connectome (SC), resulting in alteration of the flow of activation throughout the brain, which can be investigated by analyzing the functional connectome (FC).

Even though it has been shown that SC and FC are related, the complex relationship between the two has not yet been fully quantified (Abdelnour, Voss, & Raj, 2014; Honey et al., 2009; Ritter, Schirner, Mcintosh, & Jirsa, 2013). SC damage may cause an upregulation of FC in specific networks as a reorganization mechanism in the early stages of MS, which then decreases in the later stages of the disease (Faivre et al., 2012; Hawellek, Hipp, Lewis, Corbetta, & Engel, 2011). Therefore, this compensatory FC may be a biomarker used to identify the variability in impairment, even if their SC damage may be similar. It is imperative to use both modalities to understand MS because of the intricate interplay between SC and FC in damage and recovery (Damoiseaux & Greicius, 2009). Several studies have shown a decrease in SC and decrease or increase of FC in particular networks are associated with motor and cognitive impairment in pwMS (Basile et al., 2014; Faivre et al., 2012; Filippi et al., 2015; A. F. Kuceyeski et al., 2015; A. Kuceyeski et al., 2018; M. A. Rocca et al., 2012). SC and FC were also used separately or together to identify differences between pwMS and healthy controls (HC), to identify differences between MS phenotypes, and to classify pwMS by upper limb impairment severity (Kocevar et al., 2016; Richiardi et al., 2012; Saccà et al., 2018; Zhong et al., 2017; Zurita et al., 2018).

An individual’s FC is usually obtained by correlating regional Blood Oxygenation Level Dependent (BOLD) signals acquired over the entire functional MRI (fMRI) scan; however, this “static” FC derivation does not consider the fluctuations in brain network topology that can occur over time (Biswal, Zerrin Yetkin, Haughton, & Hyde, 1995; Damaraju et al., 2014). Dynamic FC (dFC) analysis is a technique that allows assessment of the topology of FCs over time via sliding windows (Allen et al., 2014). There is increased interest in using dFC to investigate pathological mechanisms in schizophrenia, bipolar, and major depressive disorders (Damaraju et al., 2014; Mennigen et al., 2018; Rashid et al., 2016; Sambataro et al., 2017). In MS, recent studies have used dFC to 1) compare clinically isolated syndrome (CIS) patients to HC, 2) analyze relationships with information processing speed in relapsing-remitting (RR) pwMS, and 3) classify cognitively impaired vs preserved pwMS (d’Ambrosio et al., 2019; Eijlers et al., 2019; Leonardi et al., 2013; Maria A. Rocca et al., 2019; Q. van Geest et al., 2018; Quinten van Geest et al., 2018).

The use of a multivariate statistical modeling, including machine learning (ML) methods, can shed light on the pathophysiological and compensatory mechanisms in neurological diseases such as stroke (Tozlu et al., 2019; Tozlu et al., 2020) and MS (Saccà et al., 2018), which could, in turn, aid clinicians in making more accurate prognoses, more effective treatment decisions and, possibly, novel therapeutics. Previous studies have used various statistical methods or ML approaches applied to pwMS data, e.g. random forest (RF), support vector machine (SVM), naïve-bayes, k-nearest neighbor, and artificial neural networks, to classify pwMS vs HC (Richiardi et al., 2012; Saccà et al., 2018; Zurita et al., 2018), to distinguish pwMS according to disease phenotype (Kocevar et al., 2016; Muthuraman et al., 2016; Zhong et al., 2017), and to predict longitudinal change in impairment (Zhao et al., 2017). However, no study to date has performed a rigorous analysis of the relative contributions of multi-modal imaging data including SC, static FC and dynamic FC to classification tasks involving pwMS.

In this paper, Random Forest models were used to classify 1) HC vs pwMS and 2) pwMS according to their impairment level. First, single-modality models were constructed using only SC, FC, or dFC; the single-modality models were then combined for an ensemble prediction. We hypothesized that models including SC could best distinguish HC from pwMS, as white matter lesions impacting SC networks are a hallmark of the disease. Furthermore, we hypothesized that models containing FC and/or dFC would best distinguish impairment levels in pwMS as this modality likely is sensitive to functional compensation mechanisms that may determine impairment levels in pwMS. Overall, our goal is to better understand mechanisms of pathology and resilience in MS, knowledge which could be used to improve the accuracy of prognoses and even develop novel therapies.

## MATERIAL AND METHODS

### Subjects

This cross-sectional study involves a total of 76 pwMS or CIS (aged 45.2±11.4 years, 50 female) and 15 healthy controls (aged 43.6±8.6 years, 8 female). All subjects had demographic data collected and underwent an MRI scan (see Table 1). Participants were excluded if they had contraindications to MRI and controls were further excluded if they had ever been diagnosed or were currently on medication for a neurological or psychological disorder. The Expanded Disability Status Scale (EDSS) was used to assess impairment level in pwMS; 23 pwMS had impairment, which we defined to be EDSS ≥ 2. The pwMS included 6 patients with CIS, 61 patients with RR, 3 patients with primary progressive (PP), and 4 patients with secondary progressive (SP). Number of spinal cord lesions was estimated from the radiology report of the spinal cord MRI.

**Table 1.**
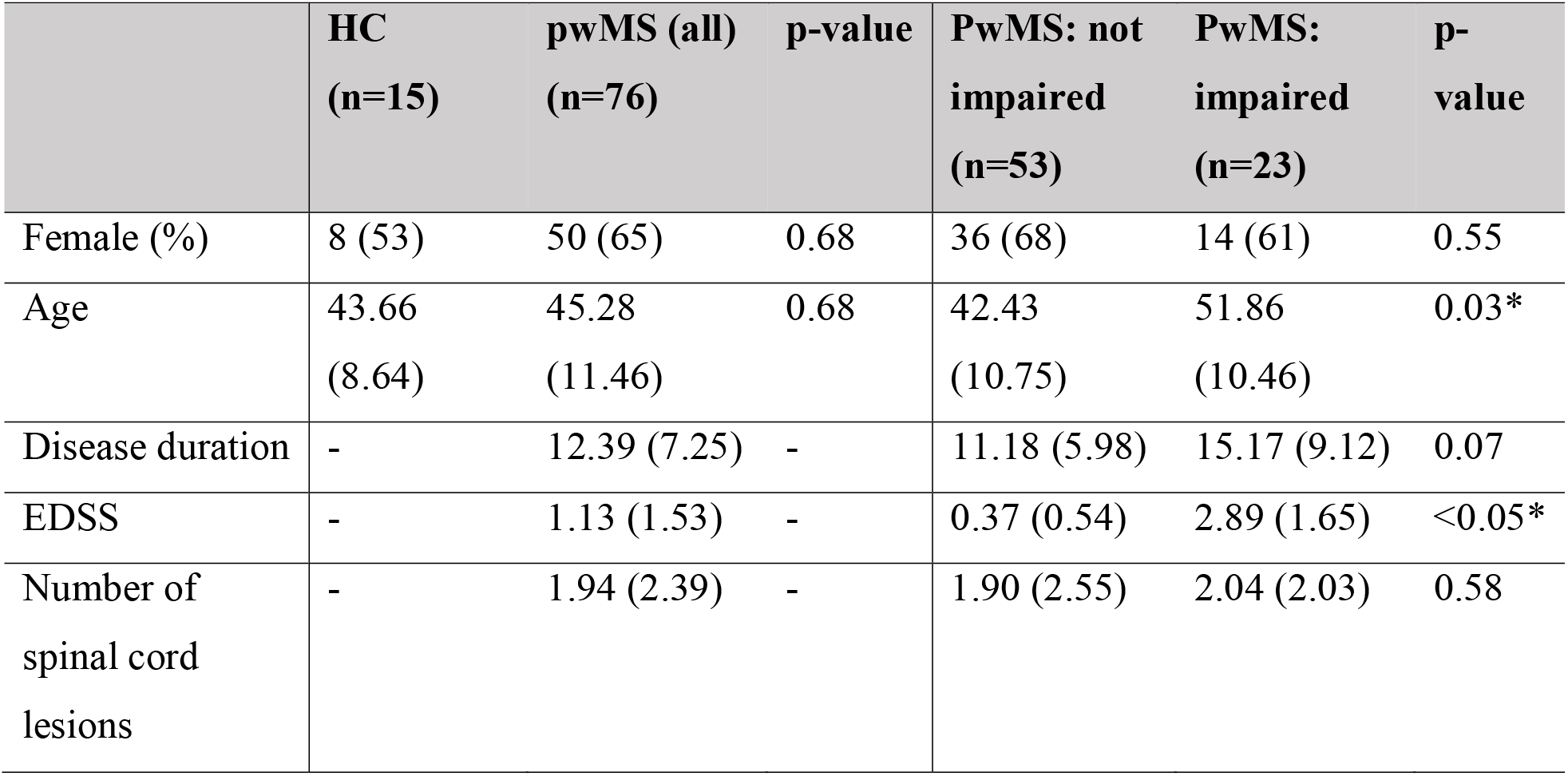
Subject demographics and clinical information. Values are presented as mean (standard deviation) for the continuous variables as the number/percent for sex. The two sets of groups were tested for differences; p-values shown are corrected for multiple comparisons.

### Image acquisition, processing and connectome extraction

MRI data were acquired on a 3T Siemens Skyra scanner (Siemens, Erlangen, Germany) with a 20-channel head-neck coil and a 32-channel spine-array coil. Anatomical MRI (T1/T2/T2-FLAIR, 1 mm3 iso-voxel), resting-state fMRI (6 min, TR = 2.3 s, 3.75 × 3.75 × 4 mm voxels) and diffusion MRI (55 directions HARDI, b=1000, 1.8 × 1.8 × 2.5 mm voxels) acquisitions were performed. Sagittal STIR images were acquired for identification of spinal lesions (TR = 3.5 s, TI=220ms, TE=45ms, in-plane resolution 0.43mm, FOV=22mm, slice thickness 3mm). Multi-echo 2D GRE fieldmaps were collected for use with both fMRI and diffusion MRI (0.75 × 0.75 × 2 mm voxels, TE1=6.69 ms, ΔTE=4.06 ms, number of TEs = 6). White matter (WM) and gray matter (GM) were segmented and GM further parcellated into 86 regions of interest (68 cortical and 18 subcortical/cerebellar) using FreeSurfer (Fischl & Dale, 2000). As described elsewhere (Kuceyeski et al., 2016), fMRI preprocessing included simultaneous nuisance regression and removal of WM and CSF effects (Hallquist, Hwang, & Luna, 2013), followed by band-pass filtering (0.008-0.09Hz) using the CONN v18b toolbox (Whitfield-Gabrieli & Nieto-Castanon, 2012) and SPM12 in Matlab. Nuisance regressors included 24 motion parameters (6 rotation and translation, temporal derivatives, and squared version of each) and the top 5 eigenvectors from eroded masks of both WM and CSF. The mean fMRI signal over all voxels in a region were calculated and the mean regional time series correlated between every pair of regions to obtain static FC.

Diffusion MRI was interpolated to isotropic 1.8mm voxels, and then corrected for eddy current, motion and EPI-distortion with the eddy command from FSL 5.0.11 (Andersson & Sotiropoulos, 2016) using the outlier detection and replacement option (Andersson, Graham, Zsoldos, & Sotiropoulos, 2016). MRtrix3Tissue (https://3Tissue.github.io), a fork of MRtrix3 (J. D. Tournier et al., 2019) was used to estimate a voxel-wise single-shell, 3-tissue constrained spherical deconvolution model (SS3T-CSD) and then compute whole-brain tractography for each subject. We performed both deterministic (sd_stream) and probabilistic (ifod2) tractography with MRtrix3 (J-D. Tournier &, F. Calamante, 2010; J-Donald Tournier, Calamante, & Connelly, 2012), after which the SIFT2 global filtering algorithm (Smith, Tournier, Calamante, & Connelly, 2015) was applied to account for bias that exists in greedy, locally-optimal streamline reconstruction algorithms. The SC matrix was constructed by taking the sum of the SIFT2 weights of streamlines connecting pairs of regions and dividing SC matrix rows by the volume of that region.

### Dynamic FC analysis

Dynamic FC matrices were calculated using a tapered, sliding window approach in the GIFT toolbox (http://mialab.mrn.org/software/gift). To quantify the reliability of results and the effect of parameter choice, Pearson’s correlation of the dFC metrics were calculated across repeated runs of varied window lengths (22, 33, 44, 55, and 66 TRs) and varied standard deviation of the Gaussian kernel (2, 3, 4, 5, and 6). Once the dFC matrices were calculated, hard-clustering and fuzzy-meta state approaches were then used to quantify FC dynamics for each subject. K-means was implemented on all dFC matrices to identify clusters of reoccurring connectivity states. The elbow criterion, calculated using Manhattan (L1) distance, was used to identify the optimal number of clusters. The following metrics were extracted from the hard-clustering analysis: 1) average dwell time in each state, or how long the individual remains in that state once they transition to it, 2) transition probability from one state to another, and 3) number of transitions from one state to another. Unlike the hard-clustering analysis that allows each dFC matrix to only be assigned to one state, the fuzzy meta state approach allows characterization of each dFC by a vector of weighted sums of different states (Miller et al., 2016). Temporal independent component analysis was used to decompose the dFCs into connectivity patterns which were maximally independent. The vectors of weights capturing the contribution of each state to a specific time window’s dFC, were discretized by assigning a non-zero integer between −4 and +4 according to its signed quartile. Each distinct weight vector is called a meta-state. The following metrics were extracted from the fuzzy meta state analysis: 1) number of distinct meta-states, 2) number of transitions from one meta state to another, 3) longest L1 distance traveled between two meta-states, and 4) total L1 distance traveled in state space.

### Mass Univariate analysis

First, demographics and clinical variables were tested for differences between the groups (HC vs pwMS and impaired vs not impaired pwMS) using Chi-squared test for qualitative variables and Wilcoxon Rank Sum test for quantitative variables. The pairwise connections in the SC and FC as well as the dFC metrics were tested for differences between the two sets of groups using the Wilcoxon rank sum test. For the SC group comparisons, only those connections present in more than 50% of the controls were tested to minimize the effect of false positives in the tractography. Differences were considered significant when p < 0.05 after Benjamini-Hochberg (BH) correction for multiple comparisons (Benjamini & Hochberg, 1995). All statistical analyses and graphs were performed using R (https://www.r-project.org), version 3.4.4 and Matlab (https://www.mathworks.com/) version R2020a.

### Classification analysis

RF models were built for two classification tasks: (1) HC vs pwMS and (2) not impaired pwMS (EDSS < 2) vs impaired pwMS (EDSS ≥ 2). Figure 1 shows the overall workflow of the study, including the three single modality models and the four ensemble models that were constructed using different combinations of the single modality models. The model was trained with outer and inner loops of k-fold cross validation (k = 5) to optimize the hyperparameters and test model performance (see Supplementary Material for details). The folds for both inner and outer loops were stratified to ensure that each fold contained the same proportion of subjects in the two classes as the original dataset. The inner loop (repeated over 10 different partitions of the training dataset only) optimized the set of hyperparameters that maximized validation AUC. A model was then fitted using the entire training dataset and those optimal hyperparameters, which was then assessed on the hold-out test set from the outer loop. The outer loop was repeated for 100 different random partitions of the data. The median and quartiles of AUC (over all 5 folds × 100 iterations = 500 test sets) were calculated to assess the performance of the methods for the single and ensemble models. Balanced accuracy, sensitivity, and specificity results were also given in Supplementary Materials to compare the results with the previous findings in the literature. A paired Wilcoxon rank sum test was used to compare classification accuracy results between models since the classification results were measured on the same test datasets. Differences were considered significant when p < 0.05 after BH correction for multiple comparisons. When the data contains class imbalance, models may tend to favor the majority class. Due to the class imbalance in our data (15 HC vs 76 pwMS and 23 impaired pwMS vs 53 not impaired pwMS), the over-sampling approach Synthetic Majority Over-sampling Technique (SMOTE) (Chawla, Bowyer, Hall, & Kegelmeyer, 2002) was used to obtain a balanced training dataset during the cross-validation. SMOTE compensates for imbalanced classes by creating synthetic examples using nearest neighbor information and has been shown to be among the most robust and accurate methods with which to control for imbalanced data (Santos, Soares, Abreu, Araujo, & Santos, 2018). The relative importance of the input variables in the final RF models was calculated using the Mean Decrease in Accuracy (MDA) metric (Breiman, 2001; Genuer, Poggi, & Tuleau-Malot, 2010), calculated by randomly permuting the value of the variable in question and assessing the drop in accuracy of the out-of-bag data. The change in accuracy is averaged over all trees and normalized by the standard deviation. The importance of the variables was averaged over all 500 models (100 outer loops × 5 models per fold).

**Figure 1:**
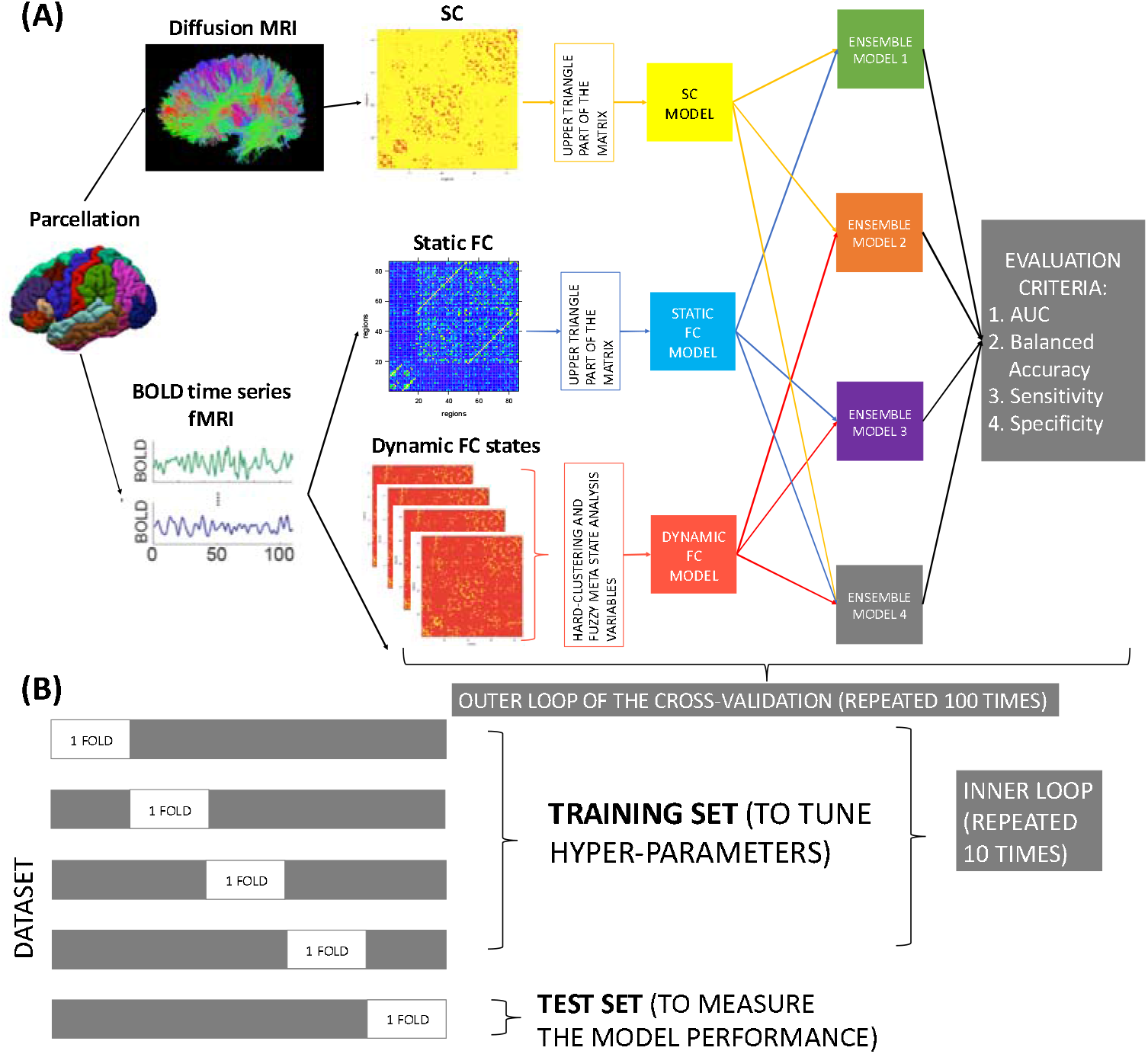
Overall workflow (A) Structural connectomes (SC), static functional connectomes (FC), and dynamic functional connectome (dFC) metrics were used to create single modality models to classify 1) HC vs pwMS and 2) pwMS according to their impairment level. The predictions from the single modality models were combined in an ensemble model approach. The classification performance of each single and ensemble model was assessed using AUC, balanced accuracy, sensitivity, and specificity. (B) The cross-validation approach that was used in the study. The original dataset was divided into 5 folds, where 4 folds were used to train the model and find best hyperparameters and one-fold was used to test the model; this procedure was performed 100 times to obtain robust estimates of model performance.

### Data availability statement

The deidentified data that support the findings of this study are available upon reasonable request. The codes that were used in this study are available. Please see https://github.com/cerent/MS-Connectome

## RESULTS

### Patient Characteristics

Table 1 shows the subjects’ demographic and clinical information including sex, age, disease duration, EDSS, and spinal cord lesion number. Demographics were not significantly different between HC and pwMS. Unsurprisingly, not impaired pwMS were younger (p = 0.03) and had a trend toward shorter disease duration (p = 0.07) but did not have significantly different sex ratio or number of spinal cord lesions.

### Mass Univariate Results

We begin by presenting the results of the mass univariate analysis that aimed to identify statistical group differences in the SC and FC matrices and in the dFC metrics between i) HC and pwMS and ii) the two impairment groups in pwMS.

#### Structural and Functional Connectome Group Comparison

We began with collecting the pair-wise connections in the upper triangular portion of the SC matrices; 666 (out of total 3655) SC pairwise connections that were non-zero in a majority of the HCs were tested for differences between groups. Forty-five SCs were significantly different between HC and pwMS, with 34 being higher in HC than pwMS. The differences between HC and pwMS is shown in Figure 2, where each significantly different connection is colored by the relative difference in medians (i.e. the difference in medians divided by sum of medians) between the groups. Unsurprisingly, the magnitude of the differences where HC > pwMS were much larger than the magnitude of the differences where pwMS > HC. The connections where HC > pwMS included those within and between subcortical, visual and somatomotor networks. The connections where pwMS > HC included mostly the left subcortical/insula to left limbic areas and from the left caudate to left thalamus and right caudate to right thalamus. There were no differences in SC between the subgroups of pwMS and no differences in FC in either group comparison.

**Figure 2:**
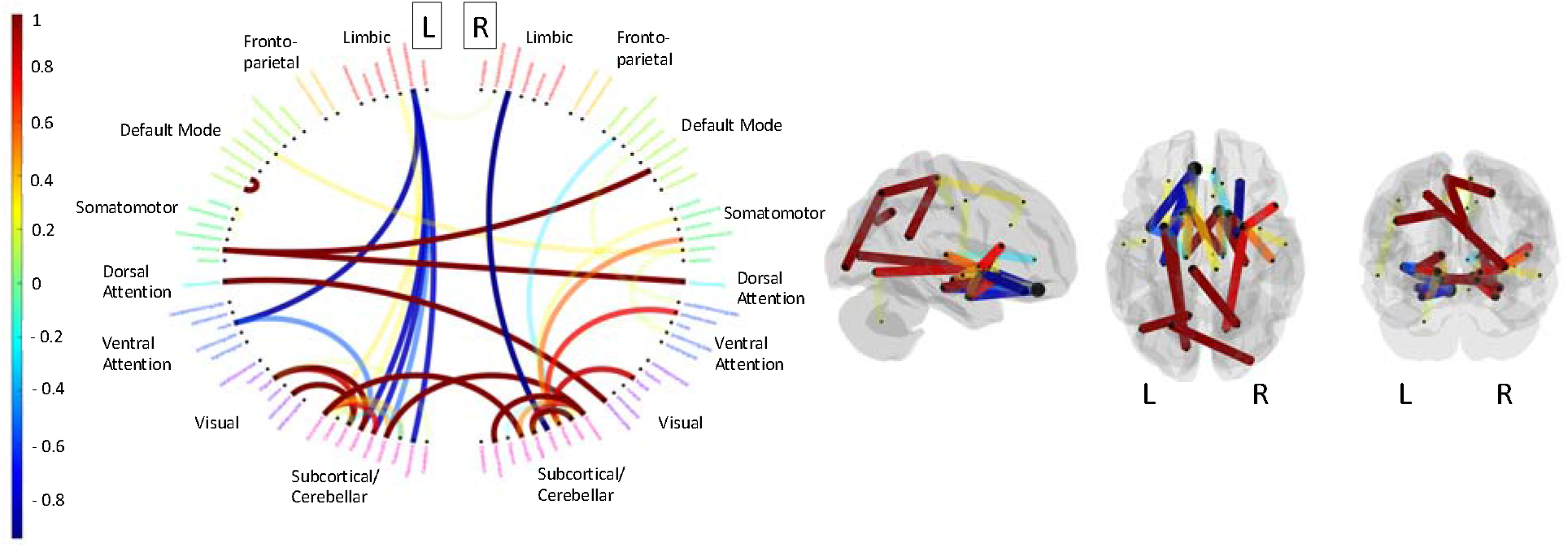
A) cluster centroids for the 4 states identified in the hard-clustering analysis of the dynamic FC. Dynamic FC metrics obtained using both (A) hard-clustering and (B) fuzzy-clustering approaches. Yellow indicates healthy controls (N = 15), medium blue all pwMS (N = 76), light blue not impaired pwMS (N = 53) and dark blue impaired pwMS (N = 23). Significant p-values (after multiple comparisons correction) are indicated by black bars and red text.

#### Dynamic FC group comparisons

For the hard-clustering analysis, the elbow curve, average dwell times and transition probabilities are given in Supplemental Figure 1; transition probabilities for each subgroup are given in Supplemental Figure 2. A window length of 22 TRs and standard deviation of 5 for the Gaussian filter resulted in the most consistent dFC measures across independent runs (see Supplementary Figure 3), therefore these parameters were used for the analyses. The four cluster centroids are shown in Figure 3A: state 1 seems to show a high within sub-network and low between subnetwork connectivity (high segregation), state 2 is characterized by large anti-correlation between the default mode network and somatomotor and dorsal/ventral attention networks, while state 4 has larger anti-correlations between the visual network and every other sub-network except for dorsal attention. Finally, state 3, the state with the largest dwell times overall, is characterized by a stronger positive correlation within and between the visual and somatomotor networks and a strong anti-correlation between those two networks and the default mode, fronto-parietal and subcortical/cerebellar networks. Figure 3 shows violin plots of the dFC metrics obtained from (B) hard-clustering and (C) fuzzy clustering approaches for the two sets of groups; significant group differences (after multiple comparison correction) are indicated. PwMS spent significantly more time in state 3 compared to HC while total distance traveled through the meta state space was significantly higher in pwMS compared to HC. There were no significant differences in any of the dFC metrics between the two subgroups of pwMS.

**Figure 3:**
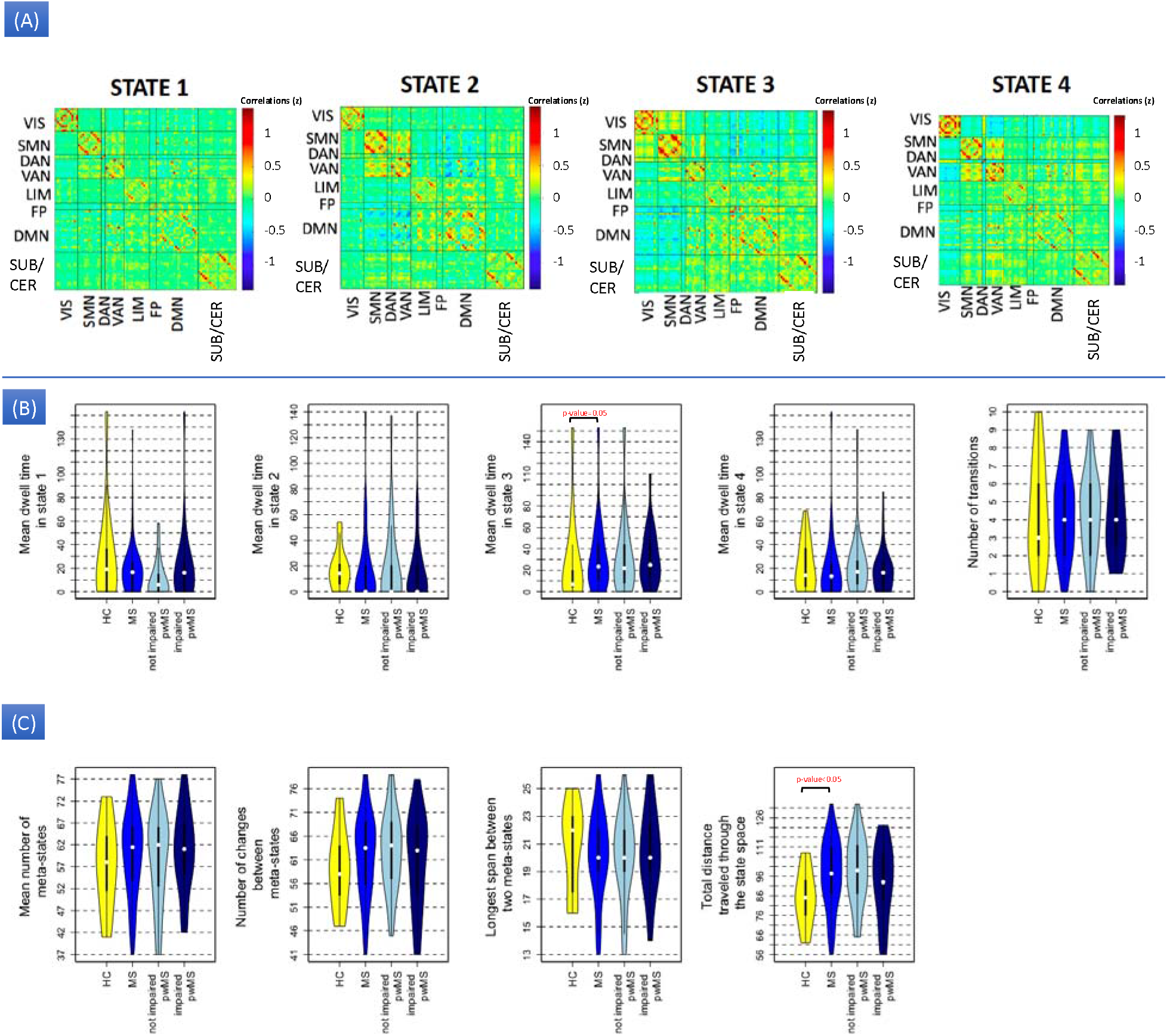
The relative difference in medians (difference divided by the sum) between the HC and MS groups for the structural connections that were significantly different between the groups (after Benjamini-Hochberg correction), where positive values (hotter colors) indicate HC > pwMS and negative values (cooler colors) indicate pwMS > HC. On the left, the circle diagram shows the name of the region clustered by functional network assignment (red = limbic, yellow = fronto-parietal, light green = default mode, dark green = somatomotor, cyan = dorsal attention, blue = ventral attention, purple = visual and pink = subcortical/cerebellar). On the right, the glass brain image shows the location of the connections within the brain, where the color of the pipes represents the difference in group medians.

### Classification Results

Here we present the assessment of the RF classification model performance and variable importance for the two classification tasks (i) HC vs pwMS and (ii) impaired vs not impaired pwMS. Figure 5 shows the distribution of AUC values (over the 500 hold-out test sets) for the single modality models based on SC, FC, and dFC separately as well as the ensemble models that combined the predictions from single models. Supplementary Table 1 contains the AUC values, balanced accuracy, sensitivity, and specificity of all the models. Unsurprisingly, the AUC for the HC vs. pwMS classification was generally higher than the AUCs for the pwMS subgroup classification. For the HC vs. pwMS task, the single-modality SC model performed significantly better than all other models, with a median AUC of 0.86. For the classification of pwMS according to their impairment level, the median AUC values ranged between 0.6 and 0.63. The single modality dFC and ensemble models that included dFC performed significantly better than the models that did not include dFC.

**Figure 4:**
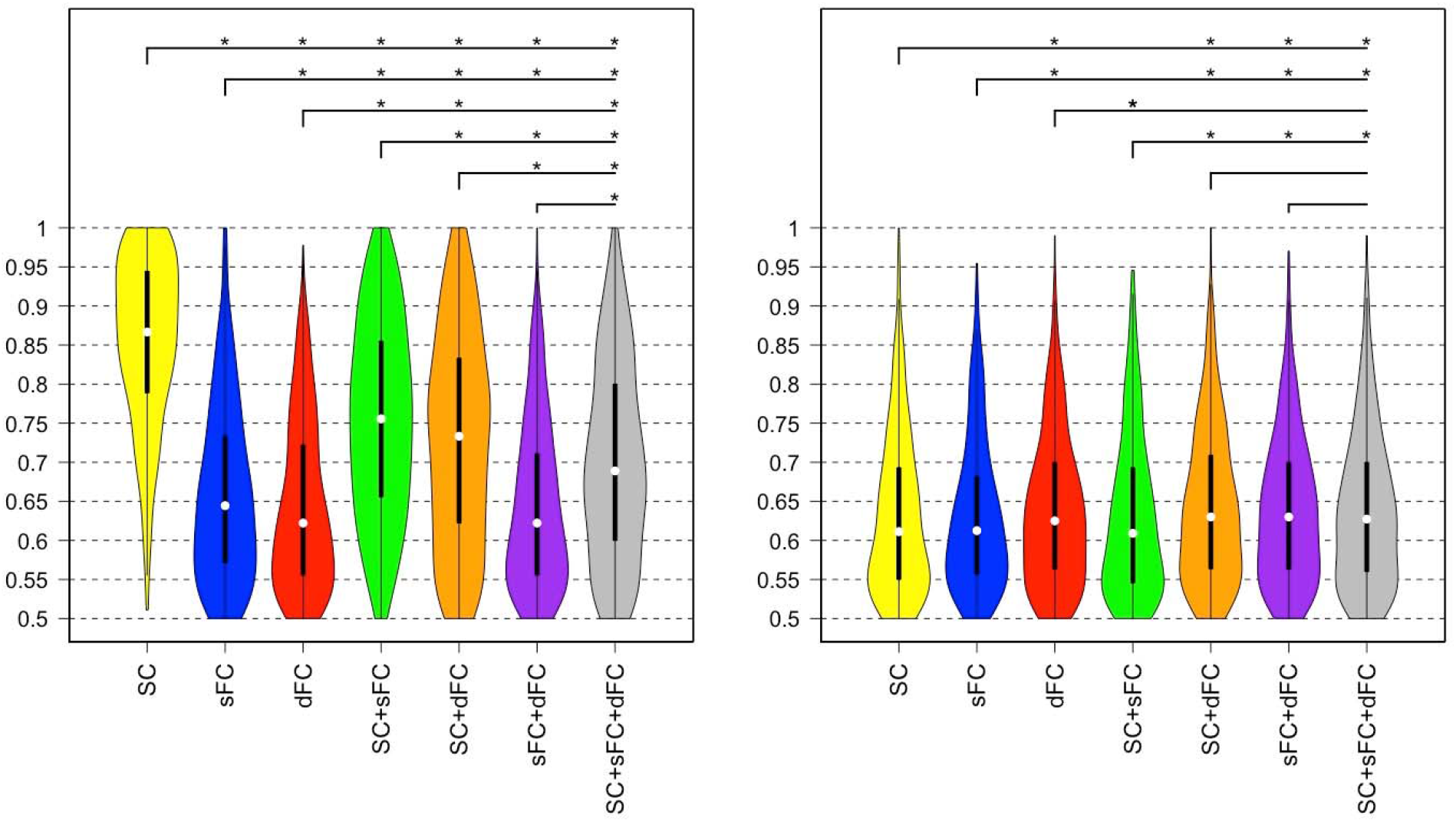
AUC results obtained from single modality and ensemble models in classifying HC vs pwMS (left panel) and pwMS according to their impairment level (right panel). The plots show the median (white dot), 1^st^ and 3^rd^ quartiles (black bar) of AUC results for single and ensemble models over the 500 hold-out test sets. The asterisks show the significant differences (p-value<0.05, BH corrected) in the distribution of AUC results between different models.

**Figure 5:**
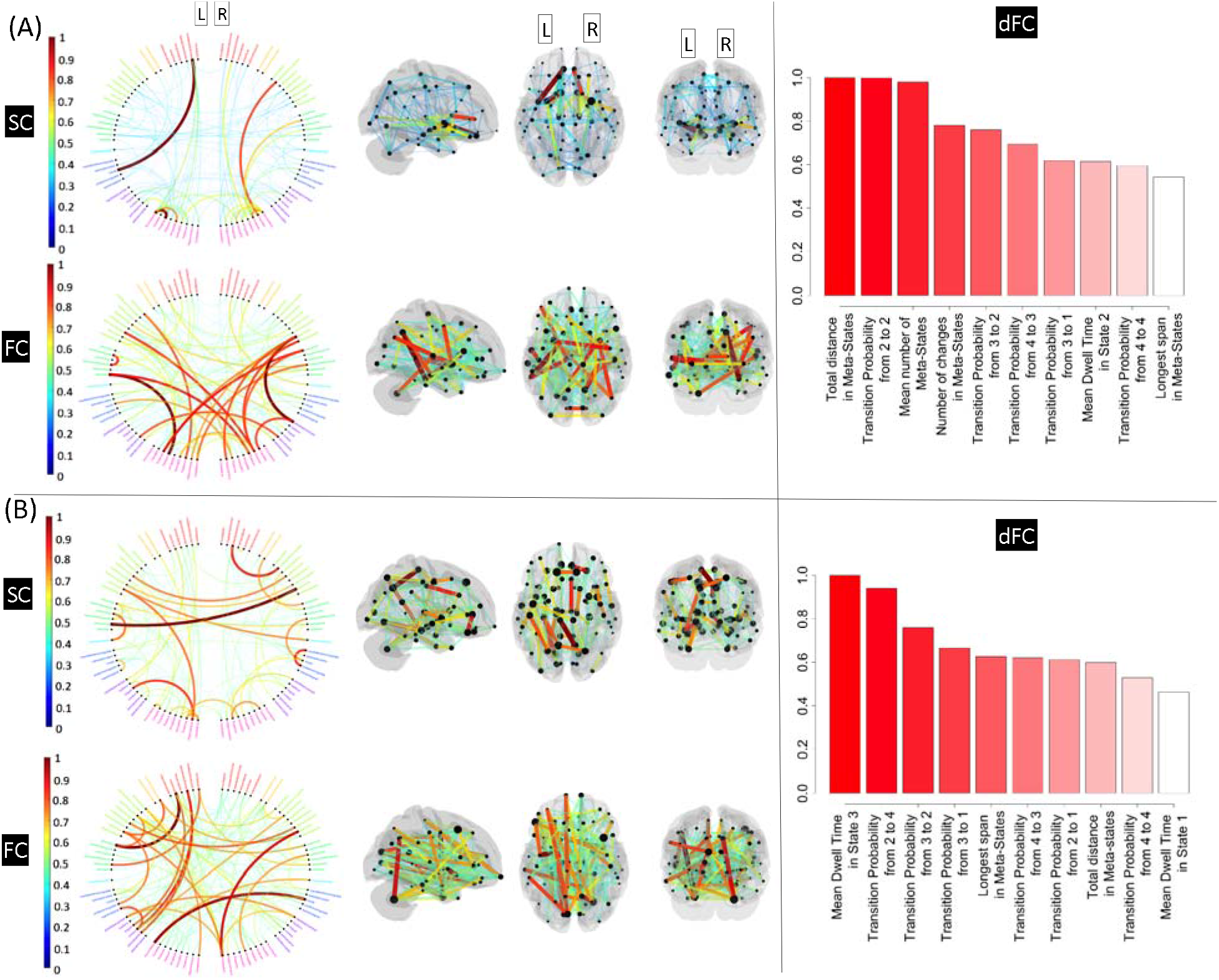
The importance of the SC, FC and dFC measures in classifying (a) HC vs pwMS and (b) pwMS according to their impairment level. Only 5% of the most important SC (top row of left panel) and FC (bottom row of left panel) are shown in the figures. Region names are provided and colored by functional network (red = limbic, yellow = fronto-parietal, light green = default mode, dark green = somatomotor, cyan = dorsal attention, blue = ventral attention, purple = visual and pink = subcortical/cerebellar). The right panels are barplots indicating the variable importance of the top 10 most important dFC metrics.

#### Variable importance

The importance of the SC, FC and dFC measures in the RF models were computed to quantify the variables that had the most influence in the two classification tasks. Scaled variable importance (relative to the maximum variable importance in each model) for the SC, FC and dFC measures are depicted in Figure 6. The FC and SC variable importance plots only show the top 5% of connections for visualization purposes. In distinguishing HC from pwMS, the most discriminative SC was the one between the left medial orbitofrontal region and the left insula while SC between subcortical areas, particularly between the accumbens and putamen and somatomotor/default mode regions also ranked in the most important variables. This agreed somewhat with the mass univariate results. FC between subcortical, default mode, somatomotor and visual networks were the most important in classifying HC and pwMS. In distinguishing between the MS subgroups, the most important SCs were intrahemispheric between default model/somatomotor networks and between subcortical and visual networks. The most discriminative FCs were found between left limbic and left somatomotor and visual and left default mode networks; the right cerebellum also had prominent importance. Since the pairwise variable importance may be noisy and are difficult to visualize, the sum of the pairwise variable importance over each of the 86 regions in the atlas are shown in Figure 7. From the regional importance visualization, it is clear that the SC and FC in the bilateral putamen were the most discriminative regions for the HC vs pwMS classification as is the SC in the left medial orbito-frontal region. SC in the left thalamus, bilateral putamen and left ventral diencephalon had the highest importance, while FC in the right cerebellum, left entorhinal, and left pars triangularis were the most discriminative predictors in separating impaired pwMS from not impaired pwMS.

**Figure 6:**
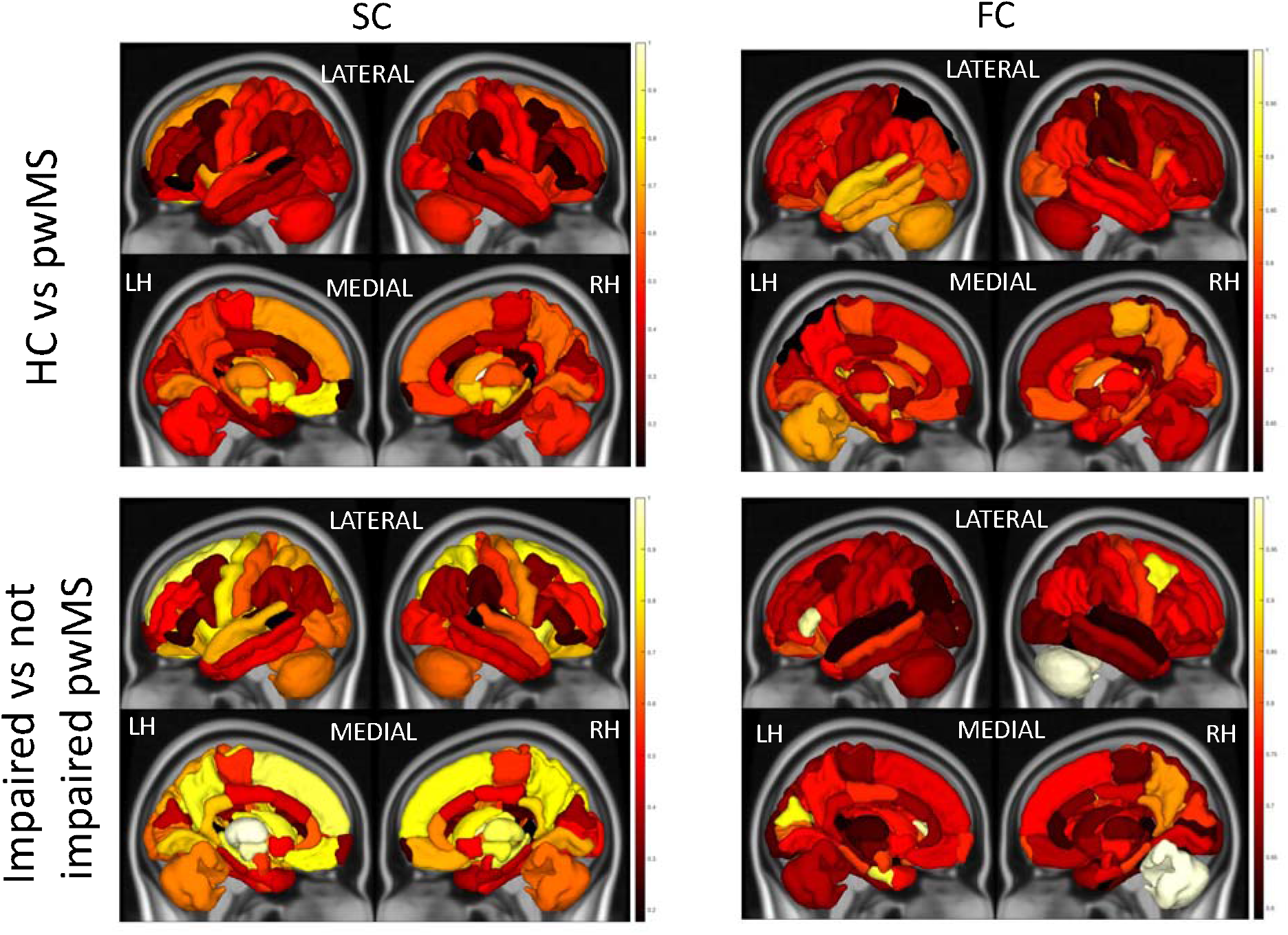
Sum of the pairwise structural and functional connections’ variable importances for each region in classifying HC vs pwMS (first row) and pwMS according to their impairment level (second row). Regional importance values are scaled by the maximum regional variable importance within each classification model.

Total distance traveled through meta states was the most discriminative dFC metric in classifying HC vs pwMS, while the average dwell time in state 3 (from the hard-clustering analysis) was the most important predictor in classifying pwMS with vs without impairment (right column Figure 6). All four of the fuzzy meta-states metrics were in the top most important metrics in classifying HC vs pwMS, as were some transition probabilities between states. In addition to various transition probabilities and mean dwell time in state 1, two meta-state measurements (the longest distance between two states and total distance traveled in state space) were also found to be highly discriminative in classifying pwMS according to their impairment level.

## Discussion

Our main findings were that, compared to HC, pwMS had weaker SC between visual, somatomotor and subcortical areas, but also spent more time in a dynamic state characterized by stronger FC between and within these same networks. We interpret this result as possibly revealing evidence for a compensatory upregulation of functional connectivity to compensate for structural connectivity damage in pwMS, i.e. the “less wiring more firing” phenomena (Daselaar et al., 2015). Furthermore, we found that pwMS traveled longer distance in dynamic FC space than HC, a result that could also be interpreted in the context of upregulation of FC. Finally, we found that classification models including SC information had the highest accuracy when discriminating between HC and pwMS as a whole, but that dynamic FC better discriminated between impairment levels within pwMS. This provides further evidence that MS is characterized by damage to the SC but impairment level within pwMS may be more related to functional compensation.

Dwell time in a dFC state, characterized by strong functional correlation of visual and somatomotor networks and a strong anti-correlation between those two networks and the default mode, fronto-parietal and subcortical/cerebellar networks, was the most important variable in the classification of impairment groups in pwMS; impaired pwMS had a trend toward longer dwell times in this state. We hypothesize this state’s increased dwell time could be the result of either a pathological or compensatory upregulation of functional coordination between visual and somatomotor systems, which play an integral role in motor function, in response to disease related damage to SC within or between these networks. In fact, a post-hoc analysis revealed a significant negative correlation between dwell time in state 3 and z-score of SC degree of the right cuneus (r = −0.22, p < 0.05) and right nucleus accumbens (r = −0.25, p < 0.05), suggesting more SC damage in these regions was related to increased dwell time in state 3. We also found that pwMS had a longer total distance traveled through the meta-state space compared to HC, which we hypothesize could reflect a reorganization mechanism in response to MS-related injury. Both the dFC and SC results support the “less wiring, more firing” hypothesis (Daselaar et al., 2015) that suggests the brain compensates for damage to SC by increased FC. Finally, we found an increase of SC in pwMS compared to HC between i) left medial-orbitofrontal regions, left insula and left subcortical areas and ii) right lateral orbitofrontal and right pallidum. This agrees with a recent finding showing an increase in betweenness centrality of the left insula and right pallidum in PwMS compared to healthy controls; there, the authors suggested this could reflect reorganization in response to MS-related damage (Llufriu et al., 2017).

Previous studies used statistical methods to differentiate between pwMS and HC, and pwMS according to their impairment severity or phenotype (Ion-Mărgineanu et al., 2017; Kocevar et al., 2016; Leonardi et al., 2013; Muthuraman et al., 2016a; Richiardi et al., 2012; Saccà et al., 2018; Stamile et al., 2015; Zhao et al., 2017; Zhong et al., 2017; Zurita et al., 2018). One recent study (Richiardi et al., 2012) showed 82% sensitivity, similar to our results, in distinguishing pwMS from HC using static FC and/or lesion load. In one of the most similar studies to date in sample size, availability of multi-modal data types and nature of classification tasks, Zurita et al., (2018) showed that using SC and static FC resulted in high accuracy of 87% in classifying HC and pwMS but the accuracy of classifying pwMS according to EDSS (≤ 1.5 or > 1.5) dropped to 63%. Here, we show 70% balanced accuracy (AUC of 0.86) in pwMS vs HC and 66% balanced accuracy (AUC = 0.63) in classifying pwMS by EDSS severity. In contrast to our results, they found that static FC was more important than SC in classifying HC and pwMS. However, their dMRI acquisition only had 15 directions compared to our higher resolution 55 directions acquisition, which likely means our SC matrices had increased sensitivity to detecting MS related damage. Zurita et al., (2018) also used SVM (as opposed to RF used here), one of the more popular approaches in discriminating pwMS vs HC using brain connectivity information (Kocevar et al., 2016; Muthuraman et al., 2016; Saccà et al., 2018; Stamile et al., 2015; Zhao et al., 2017; Zhong et al., 2017; Zurita et al., 2018). We did apply SVM with a non-linear kernel on our dataset; there were no significant differences in performance compared to our RF approach.

The most discriminative pairwise SCs in distinguishing HC from pwMS were found between the limbic/default model/somatomotor networks and subcortical areas. Particularly important was the connection from medial orbito-frontal to insula, which was one of the connections showing an increase in pwMS compared to HC in the univariate analysis. At a regional level, the SC of the bilateral putamen played the largest role in discriminating between pwMS and HC while these regions as well as the left thalamus and left ventral diencephalon played the largest role in discriminating pwMS by impairment level. The putamen and thalamus play an important role in movement, motor function and cognition (Batista et al., 2012; Henry et al., 2008) and are known to be among the first and most affected regions in MS (Bergsland et al., 2012; Santiago, Guàrdia, Casado, Carmona, & Arbizu, 2007). It has been shown that structural changes (i.e. volume loss) in the putamen and thalamus was associated with the transition from CIS to clinically definite MS and motor/cognitive impairment in pwMS (Batista et al., 2012; Bergsland et al., 2012; Calabrese et al., 2011; Ramasamy et al., 2009). Previous work in our group has shown the thalamus and putamen to be the only regions with relationships between increased atrophy and more abnormalities in connecting white matter in pwMS (A. F. Kuceyeski et al., 2015), which could indicate an impairment-relevant increase in vulnerability of these deep gray matter structures to the white matter damage that is the hallmark of MS.

Previous studies have shown that MS impacts FC in temporal, parietal and subcortical structures (Liu et al., 2016; Richiardi et al., 2012). Our findings showed that the most discriminative pairwise FCs in distinguishing pwMS from HC were between default-mode/somatomotor/visual regions and subcortical areas, and the most discriminative pair-wise FCs in distinguishing pwMS by impairment were between (primarily left) default-mode/somatomotor/visual to limbic regions, and to/from right cerebellum. The prominence of somatomotor connections is not surprising, as motor impairment is the clinical impairment of interest in this study and, of course, FC decreases in these areas have been shown to be related to increased EDSS (Schoonheim et al., 2014); furthermore, the default-mode network has been shown to be aberrant in pwMS (Greicius et al., 2007; Kutzelnigg & Lassmann, 2005; M A Rocca et al., 2010). When the FC importance was analyzed at a region level, the left temporal lobe and left cerebellum ranked in the most important predictors in classifying HC vs pwMS, while the right cerebellum, left entorhinal cortex and right pars triangularis were the most important regions in identifying the pwMS by impairment level. The prominence of the cerebellum in both classifications is in line with previous studies linking motor/cognitive disability and altered FC in the cerebellum (Dogonowski et al., 2014; Pasqua et al., 2020).

No study to date has used the brain’s dynamic FC features to classify HC vs pwMS or pwMS by impairment level. However, one study in 50 CIS patients (47 of which converted to MS) had similar dynamic FC properties compared to controls at baseline but one of the meta-state dynamic FC measurements, the distance traveled in meta-state space, increased in CIS/pwMS over 2 years to levels above and beyond HC (Maria A. Rocca et al., 2019). In another recent study, dFC metrics were compared between i) pwMS and HC and ii) pwMS with cognitive impairment vs preservation (d’Ambrosio et al., 2019). There, they showed no differences between HC and pwMS but pwMS without cognitive impairment showed increased dynamic fluidity compared to pwMS with cognitive impairment by exhibiting longer distance traveled in meta-state space, more meta-states visited and more frequent changes between meta-states. A few limitations of that study were that the data was collected across 7 sites; the authors discuss this as having a non-negligible effect on the results. Still, both of these studies indicate that, at least early on in the disease, pwMS may compensate by increasing the dynamic range of their FC, which is in agreement with our findings showing increased distance traveled in pwMS vs HC, which was more driven by the not impaired pwMS. Along the same lines, one interventional MS study showed increased strength between sensorimotor and cognitive networks in a certain dFC state was related to training induced improvements in motor function (Cordani et al., 2019).

In our study, we also compared the classification accuracy of probabilistic and deterministic tractography approaches when constructing SC. Which of these methods more faithfully reconstructs white matter pathways, particularly in the presence of pathology, is an open question. To our knowledge, there is no other study directly comparing the two tractography approaches in terms of their classification accuracy in HC vs pwMS and/or pwMS by impairment severity. Previous studies taking one approach or the other resulted in similar accuracies (Kocevar et al., 2016; Stamile et al., 2015; Zurita et al., 2018), which is confirmed by our findings (see Supplementary Table 1 and 2). Despite the similarity in classification performance, the variable importance measures (and univariate analyses comparing SC between groups) varied across tractography methods. In fact, the correlation between pairwise SC variable importances for the deterministic and probabilistic models was significant with a correlation coefficient of 0.15 for HC vs pwMS and with a correlation coefficient of 0.11 for impaired vs not impaired pwMS.

### Limitations

The less noisy correlation for region-wise SC variable importance between the deterministic and probabilistic models was higher (r=0.39 for HC vs pwMS and r=0.22 for impaired vs not impaired pwMS). Limitations The main limitations of our study were the cross-sectional nature and size of the sample. We were restricted to inferring cross-sectional relationships of brain networks properties and disability; a more clinically applicable model would be one capable of predicting with reasonable accuracy future disability for better patient management. There were only 23 not impaired pwMS and 15 controls which limited the ability to train robust models accurate in novel data. Future work includes larger, longitudinal datasets from a similar cohort. In addition, the MRI acquisition parameters could be changed to obtain higher resolution information, including reducing the TR in the fMRI scan and increasing the number of b-values in the dMRI scan. Finally, in our dFC analysis, the BOLD time series was divided using a fixed window length; however, wavelet transforms may allow different lengths for different frequency bands and will also be explored in future studies.

### Conclusion

In conclusion, machine learning applied to multi-modal connectome metrics gave high classification performance, particularly in distinguishing pwMS from HC. SC proved to be the most discriminative modality in classifying HC vs pwMS, and pwMS exhibited weaker SC within and between subcortical, visual and somoatomotor networks. Furthermore, models including dFC metrics outperformed others in classifying pwMS into impairment severity categories; there, the most important variable was dwell time in a dFC state characterized by strong FC between and among visual and somatomotor networks. These results suggest that damage to SC, particularly in the subcortical, visual and somatomotor networks, is a hallmark of MS, and, furthermore, that increased functional coordination between these same regions may be related to severity of motor disability in MS. This upregulation of functional connectivity in structurally damaged connections within subcortical, visual and somatomotor networks may be either a pathological or compensatory mechanism in MS. The use of multi-modal connectome imaging has the potential to shed light on mechanisms of disease and compensation in MS, thus enabling more accurate prognoses and, possibly, the development of novel therapeutics.

## Acknowledgements

C.T. helped with image post-processing, carried out the statistical analyses and wrote the article.

K.J. collected the data, performed pre- and post-processing of MRI data and reviewed the article.

S.G. collected the data, helped interpret results and reviewed the article.

A.K. designed and supervised the study, collected the data and edited the article.

## Citation gender diversity statement

We used classification of gender based on the first names of the first and last authors (Dworkin et al., 2020), with possible combinations including male/male, male/female, female/male, and female/female. The gender balance of papers cited within this work was quantified using gender-api.com. The authors with a gender estimation accuracy lower than 90% were checked using manual gender determination from authors’ publicly available pronouns. Among the 65 cited works, 2 articles had only one author. Among the 64 cited works with more than one author, 40% (n = 26) were MM, 34% (n = 22) were WM, 16% (n = 10) were MW, and 10% (n = 6) were WW.

## Abbreviations

AUC: Area Under Receiver Operating Characteristic Curve
BOLD: Blood Oxygenation Level Dependent
CART: Classification and Regression Trees
ChaCo: Change in Connectivity
CI: Cognitively impaired
CIS: Clinically Isolated Syndrome
CP: Cognitively preserved
EDSS: Expanded Disability Status Scale
EN: Elastic-Net
EM: Ensemble Model
GI: Gini Index
HC: Healthy controls
LR: Logistic Regression Negative Predictive
Value: NPV Positive Predictive
Value: PPV
PP: Primary progressive
RMSE: Root of Mean Squared Error
RF: Random Forest
rs fMRI: Resting state functional MRI
RR: Relapsing remitting
SP: Secondary progressive
SVM: Support Vector Machine

## References

Abdelnour, F., Voss, H. U., & Raj, A. (2014). Network diffusion accurately models the relationship between structural and functional brain connectivity networks. NeuroImage, 90, 335–347. https://doi.org/10.1016/j.neuroimage.2013.12.039

Allen, E. A., Damaraju, E., Plis, S. M., Erhardt, E. B., Eichele, T., & Calhoun, V. D. (2014). Tracking whole-brain connectivity dynamics in the resting state. Cerebral Cortex, 24(3), 663–676. https://doi.org/10.1093/cercor/bhs352

Andersson, J. L. R., Graham, M. S., Zsoldos, E., & Sotiropoulos, S. N. (2016). Incorporating outlier detection and replacement into a non-parametric framework for movement and distortion correction of diffusion MR images. NeuroImage, 141, 556–572. https://doi.org/10.1016/j.neuroimage.2016.06.058

Andersson, J. L. R., & Sotiropoulos, S. N. (2016). An integrated approach to correction for off-resonance effects and subject movement in diffusion MR imaging. NeuroImage, 125, 1063–1078. https://doi.org/10.1016/j.neuroimage.2015.10.019

Barkhof, F. (2002). The clinico-radiological paradox in multiple sclerosis revisited. Current Opinion in Neurology, 15(3), 239–245.

Basile, B., Castelli, M., Monteleone, F., Nocentini, U., Caltagirone, C., Centonze, D., … Bozzali, M. (2014). Functional connectivity changes within specific networks parallel the clinical evolution of multiple sclerosis. Multiple Sclerosis Journal, 20(8), 1050–1057. https://doi.org/10.1177/1352458513515082

Batista, S., Zivadinov, R., Hoogs, M., Bergsland, N., Heininen-Brown, M., Dwyer, M. G., … Benedict, R. H. B. (2012). Basal ganglia, thalamus and neocortical atrophy predicting slowed cognitive processing in multiple sclerosis. Journal of Neurology, 259(1), 139–146. https://doi.org/10.1007/s00415-011-6147-1

Benjamini, Y., & Hochberg, Y. (1995). Controlling the False Discovery Rate: A Practical and Powerful Approach to Multiple Testing. Journal of the Royal Statistical Society: Series B (Methodological), 57(1), 289–300. https://doi.org/10.1111/j.2517-6161.1995.tb02031.x

Bergsland, N., Horakova, D., Dwyer, M. G., Dolezal, O., Seidl, Z. K., Vaneckova, M., … Zivadinov, R. (2012). Subcortical and cortical gray matter atrophy in a large sample of patients with clinically isolated syndrome and early relapsing-remitting multiple sclerosis. American Journal of Neuroradiology. https://doi.org/10.3174/ajnr.A3086

Biswal, B., Zerrin Yetkin, F., Haughton, V. M., & Hyde, J. S. (1995). Functional connectivity in the motor cortex of resting human brain using echo-planar mri. Magnetic Resonance in Medicine, 34(4), 537–541. https://doi.org/10.1002/mrm.1910340409

Breiman, L. (2001). Random Forests. Machine Learning, 45, 5–32.

Calabrese, M., Rinaldi, F., Mattisi, I., Bernardi, V., Favaretto, A., Perini, P., & Gallo, P. (2011). The predictive value of gray matter atrophy in clinically isolated syndromes. Neurology, 77(3), 257–263. https://doi.org/10.1212/WNL.0b013e318220abd4

Chawla, N. V., Bowyer, K. W., Hall, L. O., & Kegelmeyer, W. P. (2002). SMOTE: Synthetic Minority Over-sampling Technique. Journal of Artificial Intelligence Research, 16, 321–357. https://doi.org/10.1613/jair.953

Cordani, C., Valsasina, P., Preziosa, P., Meani, A., Filippi, M., & Rocca, M. A. (2019). Action observation training promotes motor improvement and modulates functional network dynamic connectivity in multiple sclerosis. Multiple Sclerosis (Houndmills, Basingstoke, England), 1352458519887332. https://doi.org/10.1177/1352458519887332

d’Ambrosio, A., Valsasina, P., Gallo, A., De Stefano, N., Pareto, D., Barkhof, F., … Rocca, M. A. (2019). Reduced dynamics of functional connectivity and cognitive impairment in multiple sclerosis. Multiple Sclerosis Journal, 135245851983770. https://doi.org/10.1177/1352458519837707

Damaraju, E., Allen, E. A., Belger, A., Ford, J. M., McEwen, S., Mathalon, D. H., … Calhoun, V. D. (2014). Dynamic functional connectivity analysis reveals transient states of dysconnectivity in schizophrenia. NeuroImage: Clinical, 5, 298–308. https://doi.org/10.1016/j.nicl.2014.07.003

Damoiseaux, J. S., & Greicius, M. D. (2009). Greater than the sum of its parts: a review of studies combining structural connectivity and resting-state functional connectivity. Brain Structure and Function. https://doi.org/10.1007/s00429-009-0208-6

Daselaar, S. M., Iyengar, V., Davis, S. W., Eklund, K., Hayes, S. M., & Cabeza, R. E. (2015). Less wiring, more firing: Low-performing older adults compensate for impaired white matter with greater neural activity. Cerebral Cortex, 25(4), 983–990. https://doi.org/10.1093/cercor/bht289

Dogonowski, A. M., Andersen, K. W., Madsen, K. H., Sørensen, P. S., Paulson, O. B., Blinkenberg, M., & Siebner, H. R. (2014). Multiple sclerosis impairs regional functional connectivity in the cerebellum. NeuroImage: Clinical, 4, 130–138. https://doi.org/10.1016/j.nicl.2013.11.005

Eijlers, A. J. C., Wink, A. M., Meijer, K. A., Douw, L., Geurts, J. J. G., & Schoonheim, M. M. (2019). Reduced Network Dynamics on Functional MRI Signals Cognitive Impairment in Multiple Sclerosis. Radiology, 292(2), 449–457. https://doi.org/10.1148/radiol.2019182623

Faivre, A., Rico, A., Zaaraoui, W., Crespy, L., Reuter, F., Wybrecht, D., … Audoin, B. (2012). Assessing brain connectivity at rest is clinically relevant in early multiple sclerosis. Multiple Sclerosis Journal. https://doi.org/10.1177/1352458511435930

Filippi, M., Valsasina, P., Vacchi, L., Leavitt, V., Comi, G., Falini, A., & Rocca, M. (2015). Consistent Decreased Functional Connectivity Among the Main Cortical and Subcortical Functional Networks in MS: Relationship With Disability and Cognitive Impairment (P6.133). Neurology, 84(14 Supplement).

Fischl, B., & Dale, A. M. (2000). Measuring the thickness of the human cerebral cortex from magnetic resonance images. Proceedings of the National Academy of Sciences of the United States of America, 97(20), 11050–11055. https://doi.org/10.1073/pnas.200033797

Genuer, R., Poggi, J.-M., & Tuleau-Malot, C. (2010). Variable Selection using Random Forests. Pattern Recognition Letters, 31(14), 2225–2236.

Greicius, M. D., Flores, B. H., Menon, V., Glover, G. H., Solvason, H. B., Kenna, H., … Schatzberg, A. F. (2007). Resting-State Functional Connectivity in Major Depression: Abnormally Increased Contributions from Subgenual Cingulate Cortex and Thalamus. Biological Psychiatry, 62(5), 429–437. https://doi.org/10.1016/j.biopsych.2006.09.020

Hallquist, M. N., Hwang, K., & Luna, B. (2013). The nuisance of nuisance regression: spectral misspecification in a common approach to resting-state fMRI preprocessing reintroduces noise and obscures functional connectivity. NeuroImage, 82, 208–225. https://doi.org/10.1016/j.neuroimage.2013.05.116

Hawellek, D. J., Hipp, J. F., Lewis, C. M., Corbetta, M., & Engel, A. K. (2011). Increased functional connectivity indicates the severity of cognitive impairment in multiple sclerosis. Proceedings of the National Academy of Sciences of the United States of America, 108(47), 19066–19071. https://doi.org/10.1073/pnas.1110024108

Henry, R. G., Shieh, M., Okuda, D. T., Evangelista, A., Gorno-Tempini, M. L., & Pelletier, D. (2008). Regional grey matter atrophy in clinically isolated syndromes at presentation. Journal of Neurology, Neurosurgery and Psychiatry, 79(11), 1236–1244. https://doi.org/10.1136/jnnp.2007.134825

Honey, C. J., Sporns, O., Cammoun, L., Gigandet, X., Thiran, J. P., Meuli, R., & Hagmann, P. (2009). Predicting human resting-state functional connectivity from structural connectivity. Proceedings of the National Academy of Sciences of the United States of America, 106(6), 2035–2040. https://doi.org/10.1073/pnas.0811168106

Ion-Mărgineanu, A., Kocevar, G., Stamile, C., Sima, D. M., Durand-Dubief, F., Van Huffel, S., & Sappey-Marinier, D. (2017). Machine Learning Approach for Classifying Multiple Sclerosis Courses by Combining Clinical Data with Lesion Loads and Magnetic Resonance Metabolic Features. Frontiers in Neuroscience, 11. https://doi.org/10.3389/fnins.2017.00398

Kocevar, G., Stamile, C., Hannoun, S., Cotton, F., Vukusic, S., Durand-Dubief, F., & Sappey-Marinier, D. (2016). Graph Theory-Based Brain Connectivity for Automatic Classification of Multiple Sclerosis Clinical Courses. Frontiers in Neuroscience, 10, 478. https://doi.org/10.3389/fnins.2016.00478

Kuceyeski, A. F., Vargas, W., Dayan, M., Monohan, E., Blackwell, C., Raj, A., … Gauthier, S. A. (2015). Modeling the relationship among gray matter atrophy, abnormalities in connecting white matter, and cognitive performance in early multiple sclerosis. American Journal of Neuroradiology, 36(4), 702–709. https://doi.org/10.3174/ajnr.A4165

Kuceyeski, A., Monohan, E., Morris, E., Fujimoto, K., Vargas, W., & Gauthier, S. A. (2018). Baseline biomarkers of connectome disruption and atrophy predict future processing speed in early multiple sclerosis. NeuroImage: Clinical, 19, 417–424. https://doi.org/10.1016/j.nicl.2018.05.003

Kuceyeski, A., Shah, S., Dyke, J. P., Bickel, S., Abdelnour, F., Schiff, N. D., … Raj, A. (2016). The application of a mathematical model linking structural and functional connectomes in severe brain injury. NeuroImage: Clinical, 11, 635–647. https://doi.org/10.1016/j.nicl.2016.04.006

Kutzelnigg, A., & Lassmann, H. (2005). Cortical lesions and brain atrophy in MS. In Journal of the Neurological Sciences (Vol. 233, pp. 55–59). Elsevier. https://doi.org/10.1016/j.jns.2005.03.027

Leonardi, N., Richiardi, J., Gschwind, M., Simioni, S., Annoni, J. M., Schluep, M., … Van De Ville, D. (2013). Principal components of functional connectivity: A new approach to study dynamic brain connectivity during rest. NeuroImage, 83, 937–950. https://doi.org/10.1016/j.neuroimage.2013.07.019

Liu, Y., Dai, Z., Duan, Y., Huang, J., Ren, Z., Liu, Z., … Li, K. (2016). Whole brain functional connectivity in clinically isolated syndrome without conventional brain MRI lesions. European Radiology, 26(9), 2982–2991. https://doi.org/10.1007/s00330-015-4147-8

Llufriu, S., Martinez-Heras, E., Solana, E., Sola-Valls, N., Sepulveda, M., Blanco, Y., … Saiz, A. (2017). Structural networks involved in attention and executive functions in multiple sclerosis. NeuroImage: Clinical. https://doi.org/10.1016/j.nicl.2016.11.026

Mennigen, E., Miller, R. L., Rashid, B., Fryer, S. L., Loewy, R. L., Stuart, B. K., … Calhoun, V. D. (2018). Reduced higher-dimensional resting state fMRI dynamism in clinical high-risk individuals for schizophrenia identified by meta-state analysis. Schizophrenia Research, 201, 217–223. https://doi.org/10.1016/j.schres.2018.06.007

Miller, R. L., Yaesoubi, M., Turner, J. A., Mathalon, D., Preda, A., Pearlson, G., … Calhoun, V. D. (2016). Higher Dimensional Meta-State Analysis Reveals Reduced Resting fMRI Connectivity Dynamism in Schizophrenia Patients. PLOS ONE, 11(3), e0149849. https://doi.org/10.1371/journal.pone.0149849

Muthuraman, M., Fleischer, V., Kolber, P., Luessi, F., Zipp, F., & Groppa, S. (2016). Structural Brain Network Characteristics Can Differentiate CIS from Early RRMS. Frontiers in Neuroscience, 10, 14. https://doi.org/10.3389/fnins.2016.00014

Pasqua, G., Tommasin, S., Bharti, K., Ruggieri, S., Petsas, N., Piervincenzi, C., … Pantano, P. (2020). Resting-state functional connectivity of anterior and posterior cerebellar lobes is altered in multiple sclerosis. Multiple Sclerosis Journal, 135245852092277. https://doi.org/10.1177/1352458520922770

Ramasamy, D. P., Benedict, R. H. B., Cox, J. L., Fritz, D., Abdelrahman, N., Hussein, S., … Zivadinov, R. (2009). Extent of cerebellum, subcortical and cortical atrophy in patients with MS. A case-control study. Journal of the Neurological Sciences, 282(1–2), 47–54. https://doi.org/10.1016/j.jns.2008.12.034

Rashid, B., Arbabshirani, M. R., Damaraju, E., Cetin, M. S., Miller, R., Pearlson, G. D., & Calhoun, V. D. (2016). Classification of schizophrenia and bipolar patients using static and dynamic resting-state fMRI brain connectivity. NeuroImage, 134, 645–657. https://doi.org/10.1016/j.neuroimage.2016.04.051

Richiardi, J., Gschwind, M., Simioni, S., Annoni, J.-M., Greco, B., Hagmann, P., … Van De Ville, D. (2012). Classifying minimally disabled multiple sclerosis patients from resting state functional connectivity. NeuroImage, 62(3), 2021–2033. https://doi.org/10.1016/J.NEUROIMAGE.2012.05.078

Ritter, P., Schirner, M., Mcintosh, A. R., & Jirsa, V. K. (2013). The Virtual Brain Integrates Computational Modeling and Multimodal Neuroimaging. Brain Connectivity, 3(2), 121–145. https://doi.org/10.1089/brain.2012.0120

Rocca, M. A., Valsasina, P., Martinelli, V., Misci, P., Falini, A., Comi, G., & Filippi, M. (2012). Large-scale neuronal network dysfunction in relapsing-remitting multiple sclerosis. Neurology, 79(14), 1449–1457. https://doi.org/10.1212/WNL.0b013e31826d5f10

Rocca, M A, Valsasina, P., Absinta, M., Riccitelli, G., Rodegher, M. E., Misci, P., … Filippi, M. (2010). Default-mode network dysfunction and cognitive impairment in progressive MS. Neurology, 74(16), 1252–1259. https://doi.org/10.1212/WNL.0b013e3181d9ed91

Rocca, Maria A., Hidalgo de La Cruz, M., Valsasina, P., Mesaros, S., Martinovic, V., Ivanovic, J., … Filippi, M. (2019). Two-year dynamic functional network connectivity in clinically isolated syndrome. Multiple Sclerosis Journal, 1352458519837704. https://doi.org/10.1177/1352458519837704

Saccà, V., Sarica, A., Novellino, F., Barone, S., Tallarico, T., Filippelli, E., … Quattrone, A. (2018). Evaluation of machine learning algorithms performance for the prediction of early multiple sclerosis from resting-state FMRI connectivity data. Brain Imaging and Behavior. https://doi.org/10.1007/s11682-018-9926-9

Sambataro, F., Visintin, E., Doerig, N., Brakowski, J., Holtforth, M. G., Seifritz, E., & Spinelli, S. (2017). Altered dynamics of brain connectivity in major depressive disorder at-rest and during task performance. Psychiatry Research - Neuroimaging, 259, 1–9. https://doi.org/10.1016/j.pscychresns.2016.11.001

Santiago, O., Guàrdia, J., Casado, V., Carmona, O., & Arbizu, T. (2007). Specificity of frontal dysfunctions in relapsing-remitting multiple sclerosis. Archives of Clinical Neuropsychology, 22(5), 623–629. https://doi.org/10.1016/j.acn.2007.04.003

Santos, M. S., Soares, J. P., Abreu, P. H., Araujo, H., & Santos, J. (2018). Cross-Validation for Imbalanced Datasets: Avoiding Overoptimistic and Overfitting Approaches [Research Frontier]. IEEE Computational Intelligence Magazine, 13(4), 59–76. https://doi.org/10.1109/MCI.2018.2866730

Schoonheim, M. M., Geurts, J. J. G., Wiebenga, O. T., De Munck, J. C., Polman, C. H., Stam, C. J., … Wink, A. M. (2014). Changes in functional network centrality underlie cognitive dysfunction and physical disability in multiple sclerosis. Multiple Sclerosis Journal, 20(8), 1058–1065. https://doi.org/10.1177/1352458513516892

Smith, R. E., Tournier, J.-D., Calamante, F., & Connelly, A. (2015). The effects of SIFT on the reproducibility and biological accuracy of the structural connectome. NeuroImage, 104(1), 253–265. https://doi.org/10.1016/j.neuroimage.2014.10.004

Stamile, C., Kocevar, G., Hannoun, S., Durand-Dubief, F., & Sappey-Marinier, D. (2015). A graph based classification method for multiple sclerosis clinical forms using support vector machine. In Lecture Notes in Computer Science (including subseries Lecture Notes in Artificial Intelligence and Lecture Notes in Bioinformatics) (Vol. 9487, pp. 57–64). Springer, Cham. https://doi.org/10.1007/978-3-319-27929-9_6

Tournier, J-D., &, F. Calamante, and a. C. (2010). Improved probabilistic streamlines tractography by 2 nd order integration over fibre orientation distributions. Ismrm.

Tournier, J-Donald, Calamante, F., & Connelly, A. (2012). MRtrix: Diffusion tractography in crossing fiber regions. International Journal of Imaging Systems and Technology, 22(1), 53–66. https://doi.org/10.1002/ima.22005

Tournier, J. D., Smith, R., Raffelt, D., Tabbara, R., Dhollander, T., Pietsch, M., … Connelly, A. (2019, November 15). MRtrix3: A fast, flexible and open software framework for medical image processing and visualisation. NeuroImage. Academic Press Inc. https://doi.org/10.1016/j.neuroimage.2019.116137

Tozlu, C., Ozenne, B., Cho, T.-H., Nighoghossian, N., Mikkelsen, I. K., Derex, L., … Maucort-Boulch, D. (2019). Comparison of classification methods for tissue outcome after ischaemic stroke. European Journal of Neuroscience, 50(10). https://doi.org/10.1111/ejn.14507

Tozlu, Ceren, Edwards, D., Boes, A., Labar, D., Tsagaris, K. Z., Silverstein, J., … Kuceyeski, A. (2020). Machine Learning Methods Predict Individual Upper-Limb Motor Impairment Following Therapy in Chronic Stroke. Neurorehabilitation and Neural Repair, 34(5), 428–439. https://doi.org/10.1177/1545968320909796

van Geest, Q., Douw, L., van ‘t Klooster, S., Leurs, C. E., Genova, H. M., Wylie, G. R., … Hulst, H. E. (2018). Information processing speed in multiple sclerosis: Relevance of default mode network dynamics. NeuroImage: Clinical, 19, 507–515. https://doi.org/10.1016/j.nicl.2018.05.015

van Geest, Quinten, Hulst, H. E., Meijer, K. A., Hoyng, L., Geurts, J. J. G., & Douw, L. (2018). The importance of hippocampal dynamic connectivity in explaining memory function in multiple sclerosis. Brain and Behavior, 8(5). https://doi.org/10.1002/brb3.954

Weinshenker, B. G., Rice, G. P. A., Noseworthy, J. H., Carriere, W., Baskerville, J., & Ebers, G. C. (1991). THE NATURAL HISTORY OF MULTIPLE SCLEROSIS: A GEOGRAPHICALLY BASED STUDY 3. MULTIVARIATE ANALYSIS OF PREDICTIVE FACTORS AND MODELS OF OUTCOME. Brain, 114, 1045–1056.

Whitfield-Gabrieli, S., & Nieto-Castanon, A. (2012). Conn: a functional connectivity toolbox for correlated and anticorrelated brain networks. Brain Connectivity, 2(3), 125–141. https://doi.org/10.1089/brain.2012.0073

Zhao, Y., Healy, B. C., Rotstein, D., Guttmann, C. R. G., Bakshi, R., Weiner, H. L., … Chitnis, T. (2017). Exploration of machine learning techniques in predicting multiple sclerosis disease course. PLOS ONE, 12(4), e0174866. https://doi.org/10.1371/journal.pone.0174866

Zhong, J., Chen, D. Q., Nantes, J. C., Holmes, S. A., Hodaie, M., & Koski, L. (2017). Combined structural and functional patterns discriminating upper limb motor disability in multiple sclerosis using multivariate approaches. Brain Imaging and Behavior, 11(3), 754–768. https://doi.org/10.1007/s11682-016-9551-4

Zurita, M., Montalba, C., Labbé, T., Cruz, J. P., Dalboni da Rocha, J., Tejos, C., … Uribe, S. (2018). Characterization of relapsing-remitting multiple sclerosis patients using support vector machine classifications of functional and diffusion MRI data. NeuroImage. Clinical, 20, 724–730. https://doi.org/10.1016/j.nicl.2018.09.002

